# Environmental and social analysis as risk factors for the spread of the novel coronavirus (SARS-CoV-2) using remote sensing, GIS and analytical hierarchy process (AHP): Case of Peru

**DOI:** 10.1101/2020.05.31.20118653

**Authors:** Edwin Badillo-Rivera, Anthony Fow-Esteves, Fernando Alata-López, Paul Virú-Vásquez, Martha Medina-Acuña

## Abstract

The novel coronavirus disease (COVID-19) generated by the SARS-CoV-2 virus was originated in the city of Wuhan (China) in December 2019, the virus began to spread in other regions of China until it spread to the rest of the world. In this research, an analysis was made of environmental factors (tropospheric column of NO_2_, vertical airflow, percentage of solid waste disposed of in open dumps and percentage of the population without any mechanism of faeces disposal) and social factors (levels of monetary poverty, percentage of the number of hospitals per population and vulnerable population) that could directly and indirectly affect the spread of SARS-CoV-2 virus in the regions of Peru. Remote sensing techniques, geographic information systems and an analysis under the multi-parametric statistical approach proposed by Saaty were used to determine which regions present greater susceptibility, vulnerability and risk of spreading the SARS-CoV-2 virus. The results show that the prevalence of high values of tropospheric NO_2_ and values close to 0 Pa/s of the vertical airflow were directly related to the number of positive cases by COVID-19. In addition, it was found that 68% of the regions of Peru are at a “high” and “very high” risk of spreading SARS-CoV-2 virus, and most of them are in northern and central Peru (Callao, Tumbes, Piura, Loreto, Lambayeque, Huancavelica, Amazonas, Cajamarca, Ucayali, Huanuco and among others), therefore, special care should be taken with the measures adopted after social isolation in order to avoid the resurgence and collapse of the of health systems. It concludes that public policies on air quality management, integrated solid waste management and sanitation services should be improved in order to reduce the risk of spreading the SARS-CoV-2 virus. This research can be replicated on a longer scale, including more variables.

**HIGHLITGHS:**

- 68% of the regions are at a “high” and “very high” risk of spreading of SARS-CoV-2
- Tropospheric NO2 concentration and number of cases of COVID-19 are related directly
- Cases of COVID-19 are linked to tropospheric NO2 and vertical airflow to 0 Pa/s
- Environmental and social factors are analyzed together in the regions of Peru

## 1. INTRODUCTION

The novel coronavirus disease (COVID-19) caused by the SARS-CoV-2 virus was originated in China, the first reported case took place in Wuhan in December 2019. Eventually, the virus started to spread in other regions of China and on January 30, 2020, the World Health Organization (WHO) declared the COVID-19 outbreak as an International Public Health Emergency (WHO, 2020a). There are not vaccine or control disease medicine available yet (Chakraborty & Maity, 2020), so the social distance is widely adopted as the principal preventive measure (Wilder-Smith & Freedman, 2020). The infection is spreading from day to day and the health system struggles to care each infected individual, particularly in countries where the infection rate has an exponential growth, such as USA, Italy, Spain, etc. (Madurai Elavarasan & Pugazhendhi, 2020). During the spreading time of the COVID-19 disease, the scientific community shows its participation worldwide with a lot of researches, such as scientific articles, reviews, short communications and preprints (W. Ahmed et al., 2020; Chen et al., 2020; Ogen, 2020; X. Sun et al., 2020; Zambrano-monserrate et al., 2020); likewise, a diversity of approaches of the impact of COVID-19 have been analyzed in different scientific fields, mainly in medicine and mental health (Lou et al., 2020; Rajkumar, 2020). Moreover, due to the situation worldwide, environmental research are increasing, specifically in the relationship between the SARS-CoV-2 virus and environmental factors such as air, water and solid waste, it is important to know that researches are still sparse, but it is worth mentioning that these research are still scarce but they warn of some consequences that not considering them in the public policies adopted by the government could bring.

Reports and short communications have been shown that the concentrations measured in the soil and from remote sensing have a significant reduction in terms of NO_2_ and PM_2.5_ (Euronews, 2020). For instance, Chen et al. (2020) reported that the NO_2_ concentration decrease by 22.8 ug/m^3^ and the PM_2.5_, by 1.4 ug/m^3^ during the quarantine in Wuhan, while, throughout China the concentration of NO_2_ decrease in 12.9 ug/m^3^ and the PM_2.5_, in 18.9 ug/m^3^, Ogen (2020) analyzed the fatality rate of COVID-19 in Germany, France, Spain and Italy using remote sensing, concluding that long exposure to the NO_2_ could be one of the main contributors to the mortality caused by COVID-19 in those regions; in addition, to other exploratory researches which analyze air quality and its relationship between the infected rate and the fatality rate of COVID-19, they are important and necessary in order to be discussed (Conticini et al., 2020; Zhu et al., 2020). In Peru, MINAM (2020a) reported that during the social isolation, the PM_2.5_ reduced its value under 10 ug/m^3^, being the lowest average in the last 3 years; however, the common level of PM_2.5_ in March, 2018 and 2019 were 75 and 44 ug/m^3^, respectively; higher than the WHO recommendation (10 ug/m^3^ to PM 2.5). Therefore, under normal conditions of industrial activities and vehicle fleet, there could be a high relationship between the number of positive cases of COVID-19 and the high level of pollution.

There are evidences that the SARS-CoV-2 virus prevails in the faeces of the infected humans. Although, its residence time in the faeces is still unknown, therefore, it represents a risk for the human health, not only for a person, but also the janitor staff and wastewater treatment plants (WWT) workers (Guan et al., 2020; Yong Zhang et al., 2020). In Latin America, only 20% of the total wastewater have treatment (Banco Mundial, 2015). In Peru, according to a report showed by INEI (2016), in the urban areas the public sewerage system coverage is 88%, in the countryside the situation is alarming because the coverage of the public sewerage system is under 18%, being the septic tank, rivers, ditch or canal the only scenario to eliminate the faeces.

Some researches point that an inadequate solid waste management could generate the spread of SARS-CoV-2 in the management cycle of the solid waste, including its workers (Nzediegwu & Chang, 2020; van Doremalen et al., 2020). In Latin America countries exist a high predominance of open dumps because it is the faster disposal method (Ziegler-Rodriguez et al., 2019); however, this method of solid waste final disposal does not take in consideration the sanitary conditions. Institutions like ACR+ (2020), ISWA (2020) and SWANA (2020) consider important to prevent the spread of COVID-19 in those situations, for this reason, countries like USA and Italy have restricted recycling and segregation programs (Zambrano-Monserrate et al., 2020), an opposite sittuation is taking place in Peru since there is a law that priorizes the recycling of industrial inorganic solid waste as an economic activity, after the social isolation (MINAM, 2020b); moreover, according to MINAM (2020) in Peru, aproximately 73% of solid waste ends up in open dumps, and it could be a way to spread the SARS-CoV-2 virus.

It is known that SARS-CoV-2 virus can survive only few hours in the environment, and it could be enough to drive the virus to other organism and change its features, so it is necessary to take multiple scenarios into account to the near future (Núñez-Delgado, 2020); until now, the analysis of the environment factors with regard to this novel disease have been studied individually and analysis that integrates social and environmental factors is necessary, whereby, the aim of this research is to analyze individually, the social and environmental factors which affects directly or indirectly the spread of SARS-CoV-2 virus using remote sensing methods, Geographic Information System and to analyze them under the deterministic multi-parametric statistical approach by Saaty (1980). Analytic Hierarchy Process (AHP) of Saaty and the perspective of the epidemiological triad (agent, host and environment) are essential to group factors and determine the regions of Peru which are susceptible to the spread of the SARS-CoV-2 virus (Méndez-Martínez et al., 2018).

## 2. MATERIALS AND METHODS

The scope of research is Peru and the analysis were carried out in the regions of Peru, that is, the 24 regions of the country and the constitutional province of Callao were considered.

### 2.1 Environmental factors

#### 2.1.1 Tropospheric NO_2_ column (TCNO_2_)

Monitoring conditions of the air quality using remote sensing is important for health researches (Putrenko & Pashynska, 2017), in large geographical areas and very fine temporal resolution (Omrani et al., 2020). NO_2_ data in the tropospheric air column was obtained from the Sentinel-5P satellite of the tropospheric monitoring instrument-TROPOMI (Eskes et al., 2019). In this research the period of typical exposure to NO_2_ long term was defined as a period of 3.5 months, that is, from January 1, 2020 to March 14, 2020, one day before the Peruvian government decreed the period of social isolation (Diario Oficial El Peruano, 2020b); furthermore, for the purpose of showing the reduction of NO_2_, the period March 16 to April 20 was analyzed. For each period, the NO_2_ average was made using the Google Earth Engine platform, these rasters were exported and worked in GIS (QGIS); where the average pixels for each region of Peru were calculated before and after quarantine.

#### 2.1.2 Vertical airflow (**VA-O**)

VA-O is an important parameter to analyze the stability of vertical airflow in the troposphere on a synoptic scale (Räisänen, 1995). For this, airflow (Pa/s) values were used to estimate the vertical velocity in positive and negative values, where the latter indicates that the airflow pattern is from bottom to top and the positive values indicate that the airflow is from top to bottom. The hypothesis that is assumed to be affirmative is that in regions where negative values of omega prevail, it will favor a better circulation and dispersion of pollutants, preventing them from remaining on the surface and taking them to a higher altitude; on the contrary, in areas where there is airflow positive value or close to 0 (atmospheric stability) will be regions where there is a greater permanence of atmospheric pollutants. The data used for airflow was products derived from Reanalysis 1 NCEP / NCAR downloaded from the NOAA / ESRL Physical Sciences Laboratory web platform (https://psl.noaa.gov/), accessed 04/30/2020), Data analyzed correspond to the monthly average airflow for the year 2019 and at a height of 850 hPa (∼ 1500 m altitude).

#### 2.1.3 *Percentage of population without any faeces disposal mechanism* **(WED)**

The information represented the number of people by region that did not present sanitation services through the public sewerage system. To date, the presence of the SARS-CoV-2 virus has not been detected in the wastewater of Peru; however, Rosa et al. (2020) mentions that the presence of the SARS-CoV-2 virus has been reported in wastewater around the world, for example, countries such as Australia, USA, France, Italy and the Netherlands (W. Ahmed et al., 2020; Medema et al., 2020; Rosa, Iaconelli, et al., 2020; F. Wu et al., 2020; Wurtzer et al., 2020). The information regarding WED was extracted from INEI (2016).

#### 2.1.4 Percentage of solid waste disposal in open dumps (SWO)

The variable reflects the percentage of the waste generated in the population and it has established environments without sanitary measures (open dumps); it should be noted that people (children and adults) work in these places doing informal recycling tasks (RPP Noticias, 2019). Peru disposes about 72% of its solid waste in the more than 1500 open dumps recorded by the Ministry of the Environment as areas degraded by solid waste (INEI, 2018b; OEFA, 2018), the percentage of solid waste disposed of in open dumps in each region of Peru was extracted from INEI (2018b).

### 2.2 Social factors

#### 2.2.1 Vulnerable population (VP)

The vulnerable population in Peru (children under 14 and adults over 60 years old) represent 38.29% of the total population (INEI, 2017), this percentage of the total population faces different obstacles (particularly infections and diseases) during critical situations, these obstacles are also related to socioeconomic conditions, according to INEI (2017), one third of children under 14 and adults over 60 years old are poor. Information on the vulnerable population was extracted from INEI (2017).

#### 2.2.2 Percentage of the number of hospitals per population (NHP)

According to INEI (2018), Peru has 19859 infrastructures in the health sector distributed in the 25 regions of Peru, including: hospitals, health centers, health posts, health institutes specialized, medical clinics and dental centers, for this research, only the data from hospitals were extracted, due to these are the main infrastructure of the health sector that welcomes people infected COVID-19 in Peru. The information was extracted on April 24, 2020 from the INEI web portal (http://m.inei.gob.pe/estadisticas/indice-tematico/health-sector-establishments/, accessed 17/05/2020). It is important to mention that for the generation of the percentage, the number of hospitals was multiplied by a factor of 100000 and it was divided by the total population registered in last national census.

#### 2.2.3 Monetary poverty (MP)

The monetary poverty data provided by INEI (2019), groups the regions of Peru into five, This data reflects that regions have a great population without the ability to acquire a basic basket of food and non-food (housing, clothing, education, health, transportation, etc.), the data was extracted from (INEI, 2019)

### 2.3 Analytic hierarchy process (AHP)

The analytic hierarchy process (AHP) is a powerful multi-criteria method introduced by Thomas Saaty, that helps to make decisions in the face of complex problems that have multiple conflicting and subjective criteria (Saaty, 1980, 1986, 1990). The AHP has been used as a tool for decision making in different fields of science, such as health (Requia et al., 2020), natural science (Lin et al., 2020) and engineering (Yin Zhang et al., 2020; Zhou & Yang, 2020). AHP begins by defining the problem to solve (the objective) and decomposing the problem into a hierarchy of decisions (Vassou et al., 2006), then a paired comparison is used between the criteria with respect to the objective, as well as among the alternatives with respect to each criterion, with a view to establishing priorities among the elements of the hierarchy. These comparisons use a scale of values from one to nine. (see **Table 1**) determined by Saaty (1980). Finally, the relative weights of the elements of each level of the hierarchical model are estimated, the value of the global priorities of the alternatives and inconsistency (coherence) are calculated (Labib, 2014).

**Table 1.** Scale of preference between two parameters in AHP Saaty (1980)

| Preference factor | Degree of preference | Explanation |
| --- | --- | --- |
| 1 | Equally | Two factors contribute equally to the objective |
| 3 | Moderately | Experience and judgment slightly to moderately favor one factor over another |
| 5 | Strongly | Experience and judgment strongly or essentially favor one factor over another. |
| 7 | Very strongly | A factor is strongly favored over another and its dominance is showed in practice. |
| 9 | Extremely | The evidence of favoring one factor over another is of the highest degree possible of an affirmation |
| 2,4,6,8 | Intermediate | Used to represent compromises between the preferences in weights 1, 3, 5, 7, and 9. |
| Reciprocals | Opposites | Used for inverse comparison. |

As a summary, **Table 2** shows all the environmental and social factors used. An individual analysis was carried out between the number of positive cases for COVID-19 in each region and factor, while in the AHP, the environmental factors were used in the susceptibility analysis and the social factors were used in the vulnerability analysis.

**Table 2.** Environmental and social factors

| Category | Factor | Variable name | Unit | Source |
| --- | --- | --- | --- | --- |
| Environmental | Tropospheric NO<sub>2</sub> column | TCNO<sub>2</sub> | mol/m<sup>2</sup> | (ESA, 2020) |
| Environmental | Vertical airflow | VA-O | Pa/s | (NOAA, 2020) |
| Environmental | Percentage of solid waste disposed in open dumps | SWO | Percentage of solid waste disposed in open dumps /region | (INEI, 2018b) |
| Environmental | Percentage of population without faeces treatment | WED | Percentage of population without faeces treatment/region | (INEI, 2016) |
| Social | Monetary poverty levels | MP | Monetary poverty levels/region | (INEI, 2019) |
| Social | Percentage of the number of hospitals per population | NHP | Percentage of the number of hospitals per population*100000/population of region | (INEI, 2018a) |
| Social | Vulnerable population | VP | Vulnerable population/region | (INEI, 2017) |
| Social | Number of positive cases to the COVID - 19* | - | Number of positive cases to the COVID - 19/region | (MINSa, 2020) |

\* Data taken as of April 28, 2020.

## 3. RESULTS

### 3.1 Environmental factors

**Fig. 2** shows the tropospheric NO_2_ distribution map before (daily average from January 1 to March 14, 2020) and after (March 16 to April 20, 2020) of the period of social isolation for the regions of Peru, while **Fig. 3** shows the vertical airflow map (Pa/s). **Fig. 4a** shows the positive cases for COVID-19, TCNO_2_ and VA-O. The greatest positive cases of COVID-19 occurred in Callao (2933 cases), Lima (20048 cases), Lambayeque (1814 cases), Piura (960 cases) and Loreto (881 cases), and they are some of these regions where observed the highest average tropospheric NO_2_ concentrations before the period of social isolation such as in Callao (41 umol /m2), Lima (16 umol /m2) and Lambayeque (10 umol /m2). In addition, in these regions were observed that vertical airflows are closer to 0. In **Fig. 4b**, It is noted that the regions that concentrated more than 40% of WED are Loreto and Ucayali; moreover, these were among the 10 regions with the highest number of positive cases due to COVID-19 nationwide, while Callao, Lima and Tacna, concentrate less than 2% of the population that does not have faeces disposal mechanisms; however, the regions of Callao and Lima showed the highest number of people affected by COVID-19. **Fig. 4b** shows that in the regions where there are a greater number of positive cases of COVID-19, they showed SWO greater than 65%, such is the case of Lima, Callao, Lambayeque, Piura, Loreto, among others. It was noted that in all the regions of Peru there is a trend of final disposal of solid waste in open dumps greater than 45%, only in Callao it is 0% because it does not have open dumps (MINAM, 2020c).

**Fig. 1.**
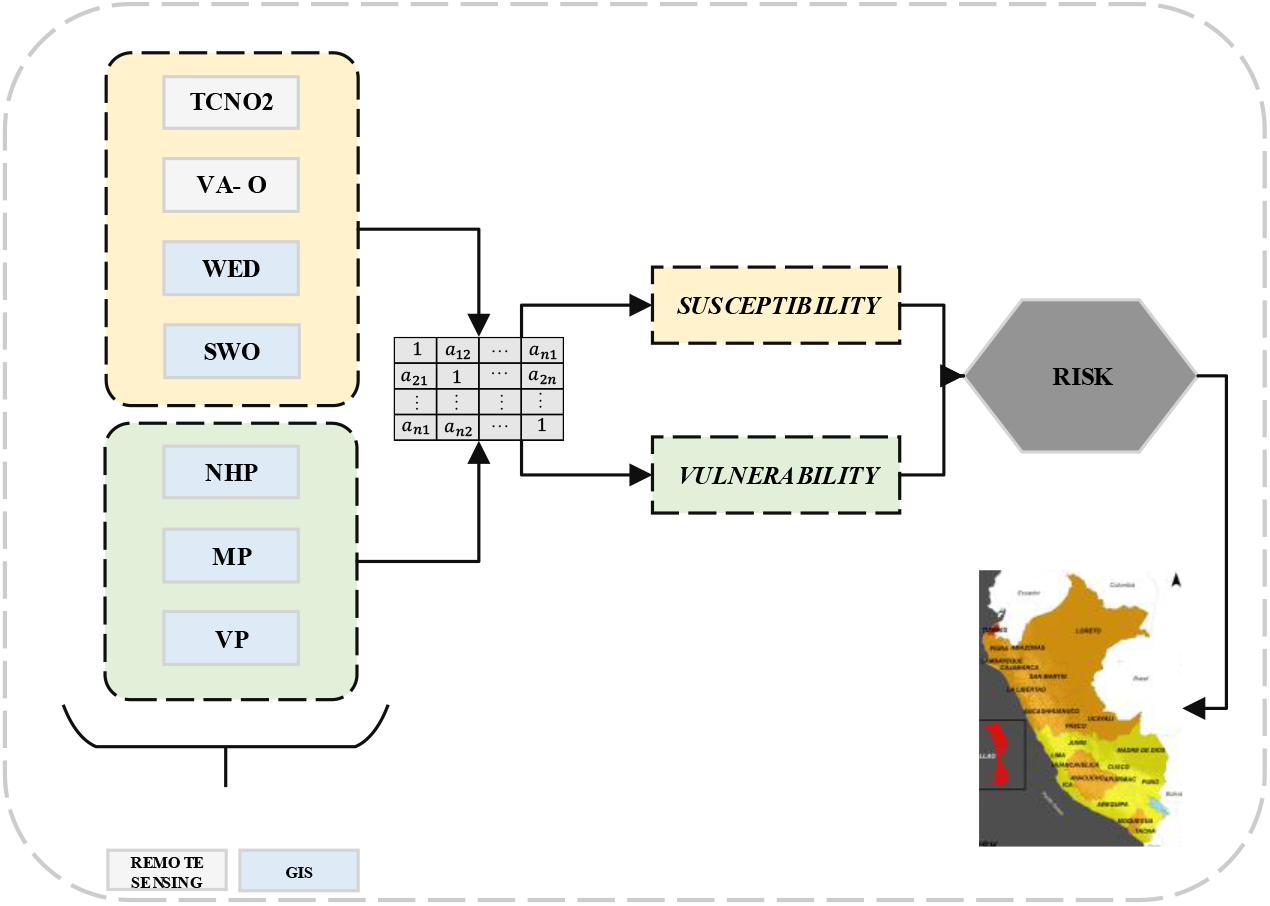
Environmental and social factors used in AHP to determine the risk of spreading SARS-CoV-2 in the regions of Peru. (TCNO_2_: Tropospheric NO_2_ column, VA-O: Vertical Airflow – Omega, WED: Percentage of population without faeces treatment, SWO: Percentage of solid waste disposed in open dumps, NHP: Percentage of the number of hospitals per population, MP: Monetary Poverty, VP: Vulnerable population)

**Fig. 2.**
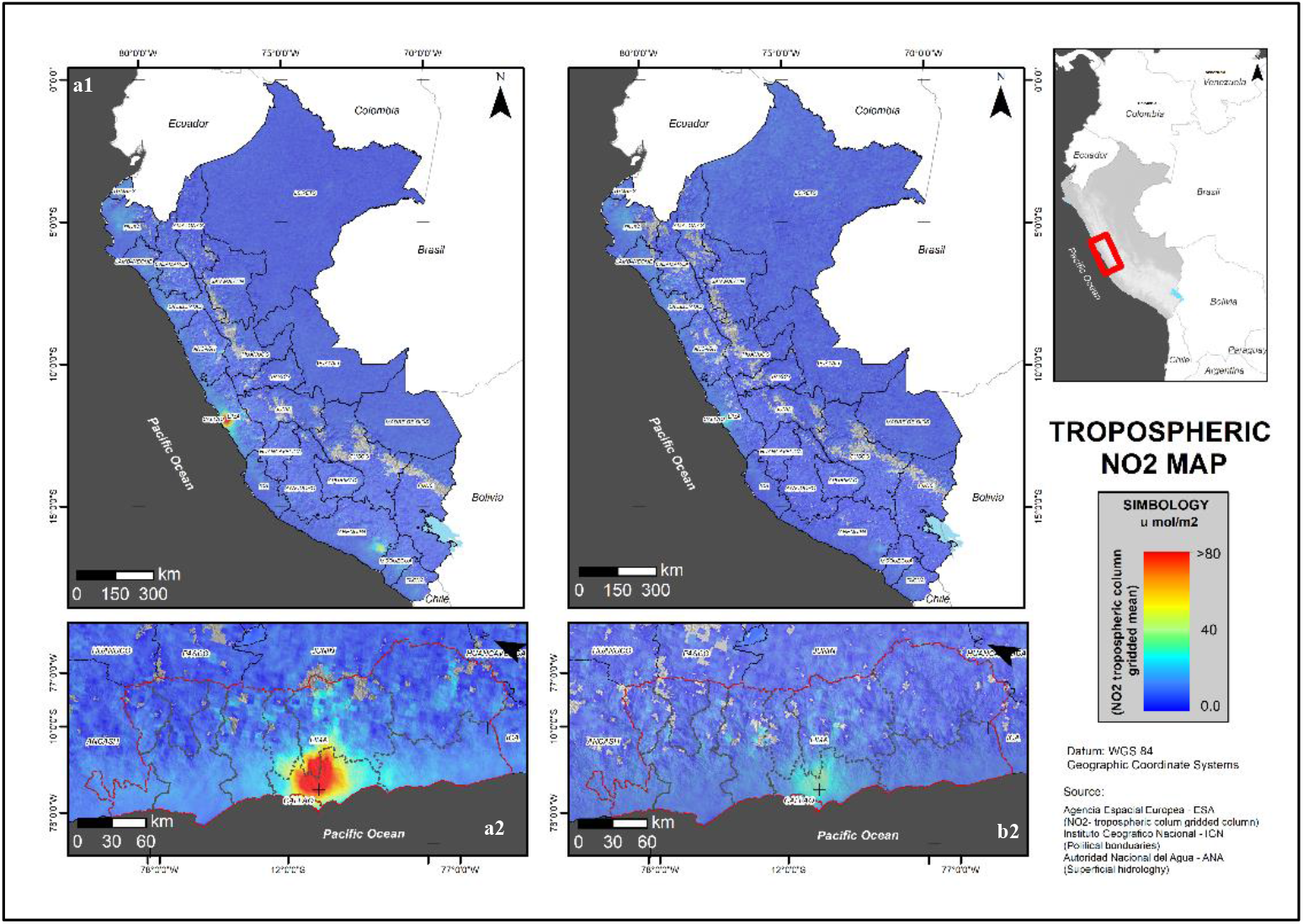
Changes in tropospheric NO_2_ concentration in the regions of Peru. The NO_2_ concentrations before the quarantine period (January 1 to March 14, 2020) are shown in **‘a1’** and **‘a2’** for Peru and the Lima region respectively, and the NO_2_ concentrations after the quarantine period (March 16 to April 20, 2020) are shown in **‘b1’** and **‘b2’** for Peru and Lima, respectively.

**Fig. 3.**
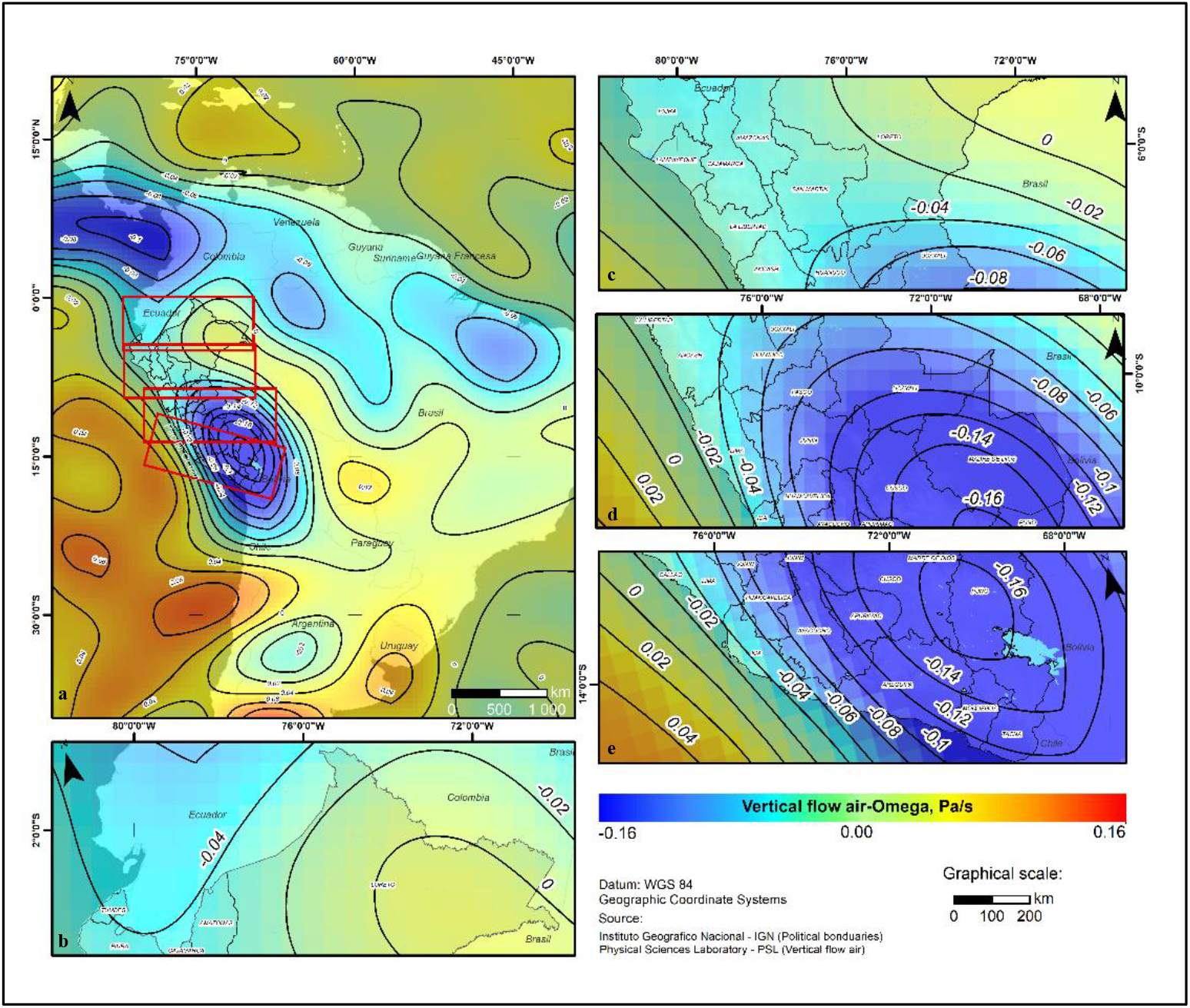
Vertical airflow (omega) (Pa/s) at 850 mb. In **‘a’** the vertical airflow for much of South America is shown, while **‘b’, ‘c’, ‘d’** and **‘e’** show an extension of the vertical airflow characteristics for the regions of Peru, in **‘b’, ‘c’** and **‘d’** values close to 0 Pa/s are observed.

**Fig. 4.**
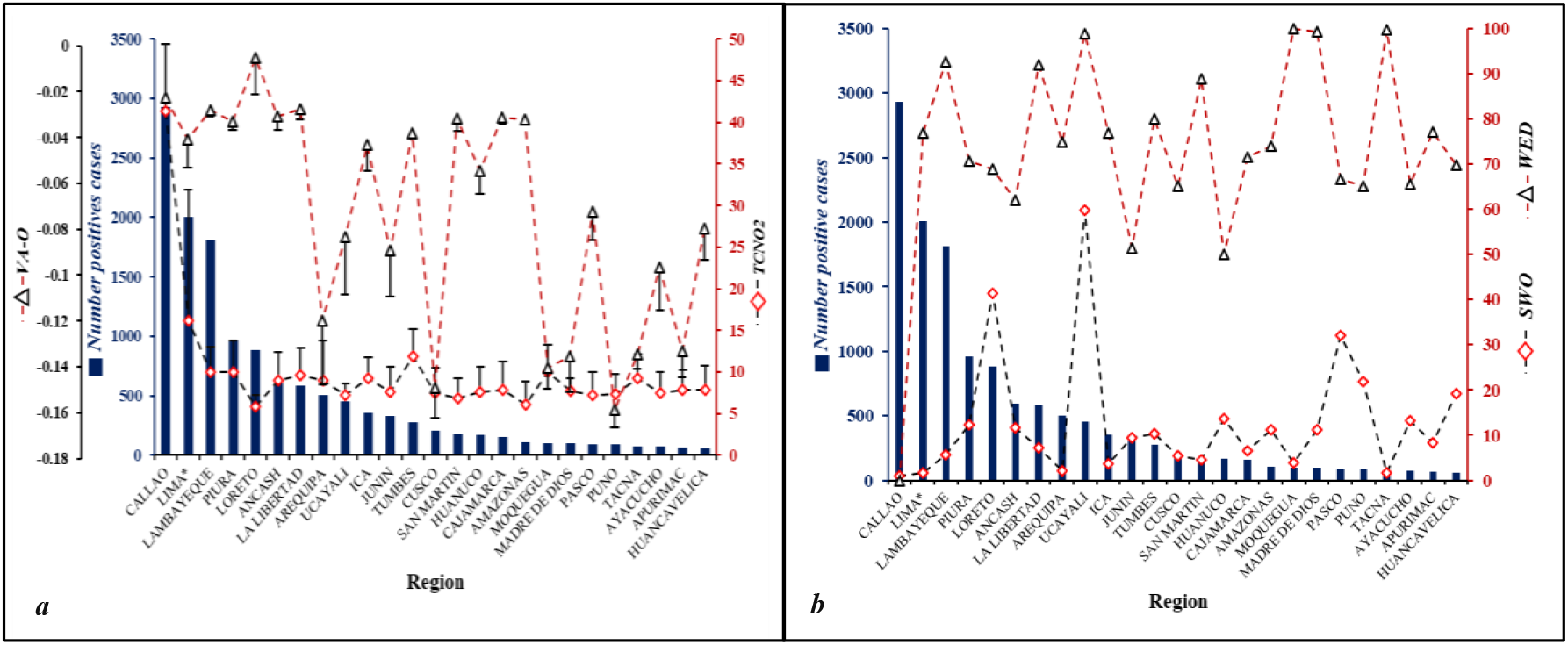
Behavior of environmental factors on the accumulation of positive cases by COVID-19 in the regions of Peru. **a**. Behavior of TCNO_2_ and VA-O on the number of infected by COVID-19; **b**. Behavior of WED and SWO on the number of infected by COVID-19. **\***The number of COVID-19 infected for the Lima region was divided by 10. (TCNO_2_: Tropospheric NO_2_ column, VA-O: Vertical Airflow – Omega, WED: Percentage of population without faeces treatment, SWO: Percentage of solid waste disposed in open dumps)

### 3.2 Social factors

**Fig. 5a** shows that on average, the percentage of hospitals per 100000 inhabitants in all regions is 1–2, even in the first 5 regions that present a greater number of positive cases for COVID-19. Regarding the vulnerable population, that is, people under 14 and over 60 years old, it is shown that in the first five regions where there are more positive cases, there are less vulnerable population. In general, it is observed that in all regions of Peru there are between 30% and 45% of vulnerable population; finally, in **Fig. 5b**, Lima, Callao, La Libertad, Piura and Loreto are the regions with the highest number of COVID-19 cases. Also, they are in group 2 to 4 of monetary poverty, on a scale of 1 to 5, where group 1 represents regions with less monetary poverty in Peru, while group 5 is the region with greater monetary poverty.

**Fig. 5.**
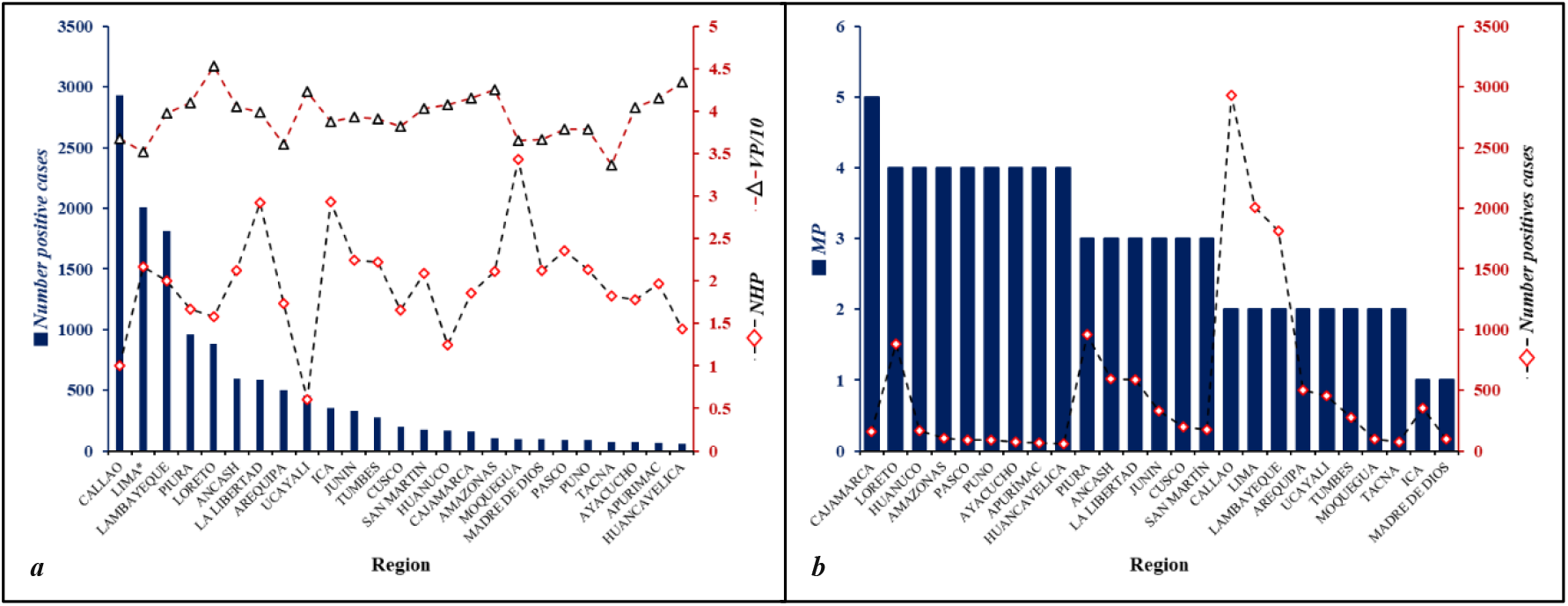
Behavior of social factors on the accumulation of COVID-19 cases in the regions of Peru. **a**. Behavior of NHP (Number and VP on the number of COVID-19 infected; **b**. Behavior of PM on the number of COVID-19 infected. **\***The number of infected by COVID-19 for the Lima region was divided by 10. (NHP: Percentage of the number of hospitals per population, MP: Monetary Poverty, VP: Vulnerable population)

### 3.3 Analytic hierarchy process

#### 3.3.1 Susceptibility factors

**Table 3** shows the relative weight values obtained from the pair-wise comparison matrix. These indicate that tropospheric NO_2_ and vertical airflow are the most important parameters with values of 0.435 and 0.407 respectively, followed by the percentage of the population without wastewater treatment with a value of 0.106; finally, the disposal of solid waste at dumps with a value of 0.052. The consistency ratio index (CR) is 0.032, value that indicates an adequate degree of consistency in the weight of each factor analyzed.

**Table 3.** Pair-wise comparison matrix, factor weights and consistency ratio of the influence of factors on susceptibility to the SAR-CoV-2 virus.

| *Influencing factors* | *Pair-wise comparison matrix* |  |  |  | *Weight* |
| --- | --- | --- | --- | --- | --- |
|  | *TCNO<sub>2</sub>* | *VA-O* | *WED* | *SWO* |  |
| TCNO<sub>2</sub> | 1.00 | 1.00 | 5.00 | 8.00 | 0.435 |
| VA-O |  | 1.00 | 5.00 | 6.00 | 0.407 |
| WED |  |  | 1.00 | 3.00 | 0.106 |
| SWO |  |  |  | 1.00 | 0.052 |
| *Consistency ratio (<0.08) = 0.032* |  |  |  |  |  |

(TCNO<sub>2</sub>: Tropospheric NO<sub>2</sub> column, VA-O: Vertical Airflow – Omega, WED: Percentage of population without faeces treatment, SWO: Percentage of solid waste disposed in open dumps)

The four environmental factors are integrated into the susceptibility index of the regions of Peru against the SAR-CoV-2 (IS_SC2_) virus that is expressed as a weighted linear sum as shown in the **Eq. (1)**.

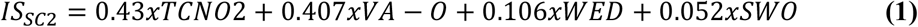

#### 3.3.2 Vulnerability factors

The values of the relative weights of the vulnerability factors analyzed obtained from the hierarchical analysis of Saaty are shown in **Table 4**, these indicate that the health facilities per population is the most important parameter 0.584, followed by the vulnerable population 0.297 that is, people under 14 and over 60 years old. Finally, monetary poverty with a value of 0.118.

**Table 4.** Pair-wise comparison matrix, factor weights and consistency ratio of the vulnerability to the SARCoV-2 virus.

| *Influencing factors* | *Pair-wise comparison matrix* |  |  | *Weight* |
| --- | --- | --- | --- | --- |
|  | *NHP* | *VP* | *MP* |  |
| NHP | 1.00 | 2.00 | 4.00 | 0.557 |
| VP |  | 1.00 | 3.00 | 0.320 |
| MP |  |  | 1.00 | 0.123 |
| *Consistency ratio ($<0.04$) = 0.017* |  |  |  |  |

(NHP: Percentage of the number of hospitals per population, MP: Monetary Poverty, VP: Vulnerable population)

In order to integrate the three factors analyzed and determine the vulnerability levels of the populations in the regions of Peru, the vulnerability index against the SAR-CoV-2 virus (IV_SC2_) was used, which is expressed as a weighted linear sum as shown in the **Eq. (2)**.

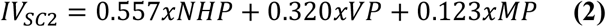

The risk levels by region against the SAR-CoV-2 virus are obtained from the product of the values of the levels of susceptibility and vulnerability (CENEPRED, 2015). **Fig. 6** shows the maps of susceptibility, vulnerability and risk at the regional level of Peru against the SAR-CoV-2 virus, the classification was made in four levels (low, medium, high and very high) according to the classification of the method (Pourghasemi et al., 2012).

**Fig. 6.**
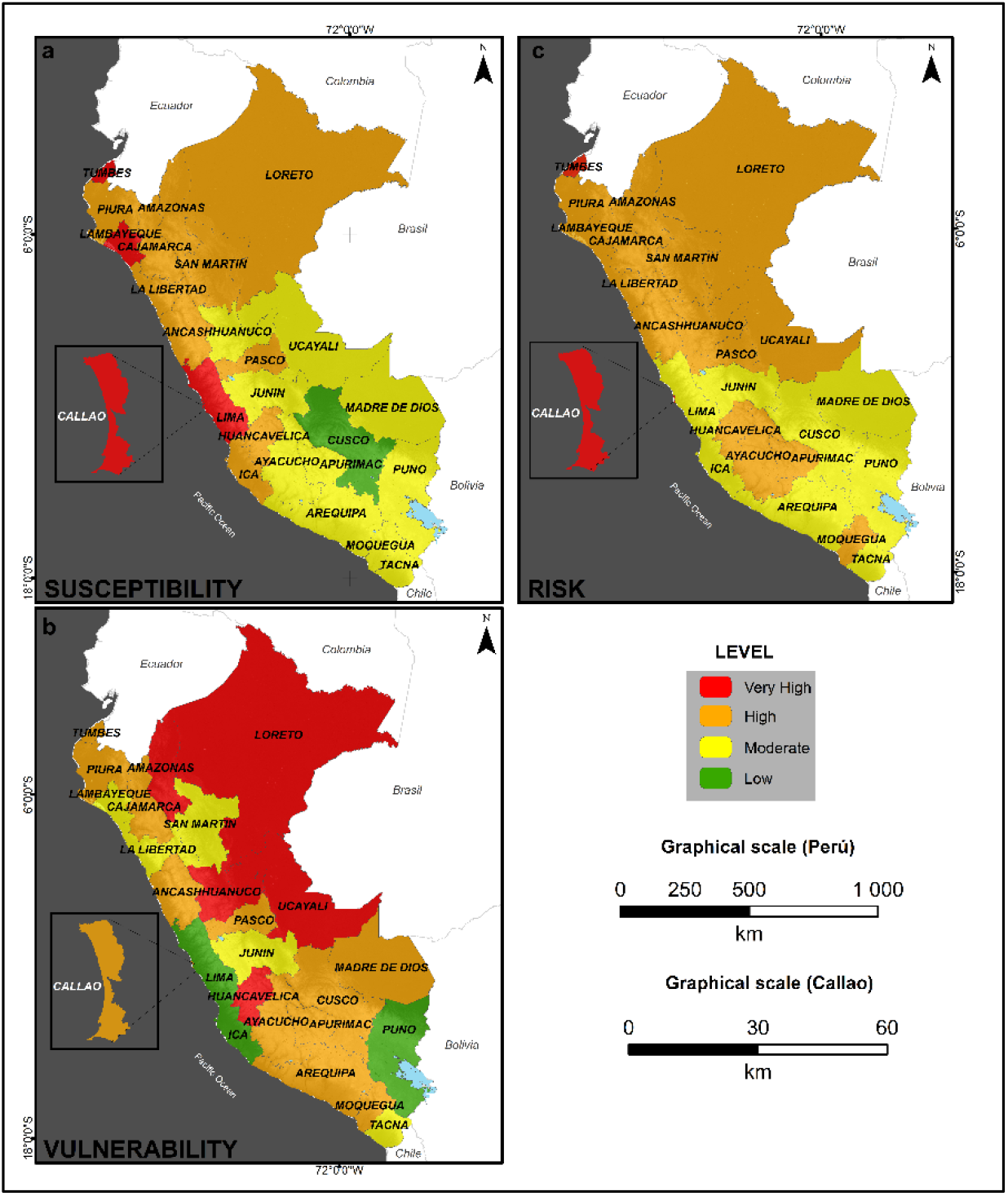
**a.** SARS-CoV-2 virus spread susceptibility map; **b**. SARS-CoV-2 virus spread vulnerability map; **c**. SARS-CoV-2 virus spread risk map.

Regions as Lambayeque, Callao, Tumbes and Lima showed a “very high” level of susceptibility to the spread of the SARS-CoV-2 virus; furthermore, La Libertad, Piura, Loreto, Ancash, Cajamarca, Amazonas, Ica, San Martin, Huancavelica and Pasco showed a “high” susceptibility level, most of these regions are located in the north and on the central coast of Peru, the other regions are at a “medium” and “low” (Cusco). It is worth mentioning that all the regions with “very high” susceptibility are found on the coast of Peru; regarding the levels of vulnerability, it was found that Huancavelica, Huanuco, Ucayali, Amazonas and Loreto showed a “very high” level of vulnerability to the spread of the SARS-CoV-2 virus. In terms of the regions that presented a level of “high” vulnerability, six of these are in southern (Moquegua, Apurimac, Cusco and Arequipa), two in the center (Pasco and Callao) and three in northern (Tumbes, Piura, Cajamarca and Ancash), the others regions are at a “medium” and “low” level of vulnerability. Regarding risk, Callao and Tumbes have a “very high” level, Piura, Loreto, Lambayeque, Huancavelica, Amazonas, Cajamarca, Ucayali, Huanuco, Ancash, Moquegua, Pasco, Ayacucho, San Martín, La Libertad and Apurimac have a level of “high”; all other regions showed a “medium” level of risk.

## 4. DISCUSSION

In the preliminary stage of the COVID-19 outbreak, a lot of research have managed to forecast in advance the dynamics of possible outbreaks using various methodologies (Hellewell et al., 2020; Ren et al., 2020; Roosa et al., 2020; K. Sun et al., 2020; Z. Wu & McGoogan, 2020). Based on that, useful evidences have been provided for the health sectors in different countries. This research is based on the individual analysis and the AHP of 8 factors that could identify those regions of Peru that are most susceptible to the risk of COVID-19 infection.

### 4.1 Environmental factors

According to TCNO_2_ over the regions of Peru, a notable reduction is observed before and after the quarantine period established by the Peruvian government of up to three times in regions with high population density, intensive industrial activity and a great activity of the automotive park such as Arequipa, Callao, Lima and Piura. The highest tropospheric NO_2_ concentration values or ‘hotspot’ are distributed in regions of the northern coasts, central coast and part of the south coast of Peru, some of them are Lima, Callao, Lambayeque and Piura, being these the regions where recorded higher positive cases for COVID-19. On the other hand, the Amazon region presents the lowest tropospheric NO_2_ concentration value, and that region is among the ten regions with the lowest number of confirmed COVID-19 cases. Therefore, it is possible that there is a direct relationship between the amount of tropospheric NO_2_ concentration and the number of confirmed COVID-19 cases (Ogen, 2020). In addition, Wu et al. (2020) researched that long-term exposure to poor air quality could exacerbate COVID-19 symptoms, and even the risk of mortality from this disease. Regarding VA-O, it is observed that its highest value (<-0.02 Pa / s), value close to 0 (which represents a stability of vertical air flow), has been detected in the Loreto region, which it is within the five regions with the highest number of confirmed COVID-19 cases. Likewise, the lowest value of VA-O has been detected in the Puno region (> –0.16 Pa/s), which is within the five regions with the lowest number of confirmed COVID-19 cases. Therefore, it is possible that there is a direct relationship between the regions that have a more stable VA-O (calm condition and favoring the concentration of pollutants) and the number of confirmed cases of COVID-19. In summary, over the regions of Peru, it founds that those with the highest number of infected by COVID-19, there is a prevalence of high tropospheric NO_2_ and vertical airflow values very close to 0 Pa/s. That is, they are areas where atmospheric stability conditions prevail, this conditions that not only tropospheric NO_2_, but also that other atmospheric pollutants are concentrated near the surface of the earth. Furthermore, Wang et al. (2020) mentions that environmental conditions can be linked to the rate of spread of the SARS-CoV-2 virus and the severity of the disease.

Regarding **WED**, regions such as Loreto and Ucayali concentrate more than 40% of their total population without faeces disposal mechanisms. According to SUNASS (2015), in both regions there are not WWT with disinfection treatment and in the regions of the Peruvian coast, where there is a greater number of positive cases for COVID-19, they have a percentage of the population without faeces treatment of less than 15%, therefore, establishing an inverse or direct relationship between the percentage of the population that does not have faeces disposal mechanisms and the number of confirmed cases of COVID-19 would not be completely adequate; however, the absence of faeces disposal mechanisms could bring public health problems related to contracting COVID-19 because there are evidences that such virus can be found in the faeces of infected people (W. Ahmed et al., 2020; Medema et al., 2020; Rosa, Iaconelli, et al., 2020; F. Wu et al., 2020; Wurtzer et al., 2020; Yong Zhang et al., 2020). Regarding the environmental factor, **SWO**, in general, in all the regions of Peru there is a trend of final disposal of solid waste in open dumps greater than 45%, only in Callao it is 0% because it does not have open dumps (MINAM, 2020c). Although a direct relationship was not found between this factor and regionally confirmed cases of COVID-19, precautions should be taken in the management of solid waste generated by people infected with COVID-19 because it could be a potential transmission path (ACR+, 2020; SWANA, 2020). In addition, van Doremalen et al. (2020) mentions that the residence time in stainless steel, plastic, cardboard and copper wastes could be linked to the number of positive cases by COVID-19, since their residence time in this type of solid waste can be several hours, therefore, the measure adopted by the Peruvian government in relation to recycling could increase the number of cases of COVID-19 infection in people involved in the solid waste management cycle (Diario Oficial El Peruano, 2020a).

### 4.2 Social factors

The **NHP** factor did not show a uniform association with the number of positive cases for COVID-19. In Peru, the health emergency due to COVID-19 has put health systems to the test; where the number of human resources in health is 1.3 per 100000 population (Diario Gestión, 2018), while OPS suggests that the number of human resources in health should be 2.3 for every 100000 population (OPS, 2015). On the other hand, WHO (2020b) indicates that when health systems are overwhelmed, morbidity is exacerbated, disability intensifies, and both mortality due to the outbreak (direct) and mortality due to previous conditions and treatable with vaccines (avoidable). Finally, the health system of Peru is closed to the limit despite the multiple efforts made by the government (El Diario, 2020). From the analysis of the results for **VP**, it was found that in all regions of Peru there is a vulnerable population between 30 and 45% of the total population, Vohora (2017) mentions that vulnerable populations are at higher risk during an adverse scenario; COVID-19 is not the exception. Lastly, it was found that the regions with the highest numbers of positive cases for COVID-19 are located in the north of Peru and correspond to monetary poverty groups 2, 3 and 4. Ahme et al. (2020) mentions that the socioeconomic disadvantages and inequalities during an epidemic become more evident. Consequently, the economic impact generated by COVID-19 in Peru has led to higher unemployment rates (M. Vinelli & A. Maurer, 2020).

### 4.3 Analytic hierarchy process

This research combined environmental and social factors analyzed under the Saaty hierarchical analysis approach, determining that the “high” and “very high” risk levels of the spread of the SARS-CoV-2 virus are found in a greater proportion in the north and central of Peru. In addition to some southern regions; These results are explained because in these regions there is a considerable level of vulnerability (evaluated based on social factors) and susceptibility (evaluated based on environmental factors). Although the model applied may be limited by the relatively small number of variables analyzed, it is the first research to analyze environmental and social factors (together) in Peru, and it is shown in a relatively simple way. Also, our results are in accordance with what was found in previous researches. For example, on May 5, 2020 the CDC – MINSA (2020) published a report indicating that 60% of the regions of Peru should strengthen case care and the control against COVID-19, among these regions are, Ancash, Callao, Lambayeque, Loreto, Piura, Tumbes, Ucayali, Lima, Ica, La Libertad, Pasco, Arequipa, Junin, Huanuco and Amazonas, while in this research it was found that 68% of the regions of Peru are at a “high” and “very high” level of risk of spreading the SARS-CoV-2 virus, in addition to the regions considered by the CDC – MINSA (2020). This research suggests that the San Martin and Ayacucho regions should be included within the prioritized regions. In the research carried out by Yaser Burhum (2020), it was shown that as of May 16, 2020, all regions maintained an R index greater than 1 (R> 1, virus in spread, R < 1, virus in disappearance), with which it can be inferred that the SARS-CoV-2 virus in Peru is still in a propagation stage, this could be related to the fact that the environmental and social factors analyzed in the present research and others not analyzed would influence the spread of the SARS virus –CoV-2.

### 4.4 Limitations

The authors acknowledge that this research has some limitations. The temporality of the data used ranges between 2015 and 2020; however, it is expected that this variation in temporality has not significantly affected the results. In Peru, to access to the geo-referenced data of positive cases by COVID-19 is not available, officially only the accumulated by regions can be obtained, this limits that the research can be carried out on a much larger scale, perhaps up to a district or focused level. In addition, the foregoing limited the addition of other variables to the model that could influence the spread of the SARS-CoV-2 virus, as has been demonstrated in Arias-Reyes et al. (2020).

## 5. CONCLUSIONS

It is concluded that the trend of environmental factors TCNO_2_ and VA-O presented a direct relation with the number of positive cases by COVID-19 in each region of Peru; however, the analyses of the other environmental and social factors show a direct relation only with some regions of Peru, so when performing the AHP the factors have complemented each other. This research is one of the few in which social and environmental factors are analyzed together, which would be associated with the spread of SARS-CoV-2 virus through a multiparametric analytical decision-making process. In relation to this, our results are linked in line with previous studies made in Peru like CDC – MINSA (2020) and Yaser Burhum (2020). We also concluded that in 68% of the regions of Peru there is a “high” and “very high” risk of spreading the SARS-CoV-2 virus due to the factors analyzed and that these are in the north of Peru. Therefore, special care should be taken after social isolation, specifically in regions as Callao, Tumbes, Piura, Loreto and Lambayeque in order to avoid a resurgence and collapse of the health systems. Based on the results presented in this research, it is considered that the Peruvian government should promote with greater force the improvement of public policies on air quality management, solid waste management and sanitation services and urban and rural sewage systems, in order to reduce the risk of spreading the SARS-CoV-2 virus. Finally, based on the results, we suggest that the methodology adopted in this research could be replicated at different scales, considering the introduction of more variables according to the reality of each research area.

## Data Availability

The data that support the plots within this paper and other findings of this study had been included in the manuscript and they are available from the corresponding authors upon reasonable request.

## ACKNOWLEDGEMENTS

The authors thank Richard Huapaya Pardavé and the research group “Sustainable Development and Environmental Research” who very cordially reviewed the scientific article. To our universities, National University of Callao and National University of Engineering.

---

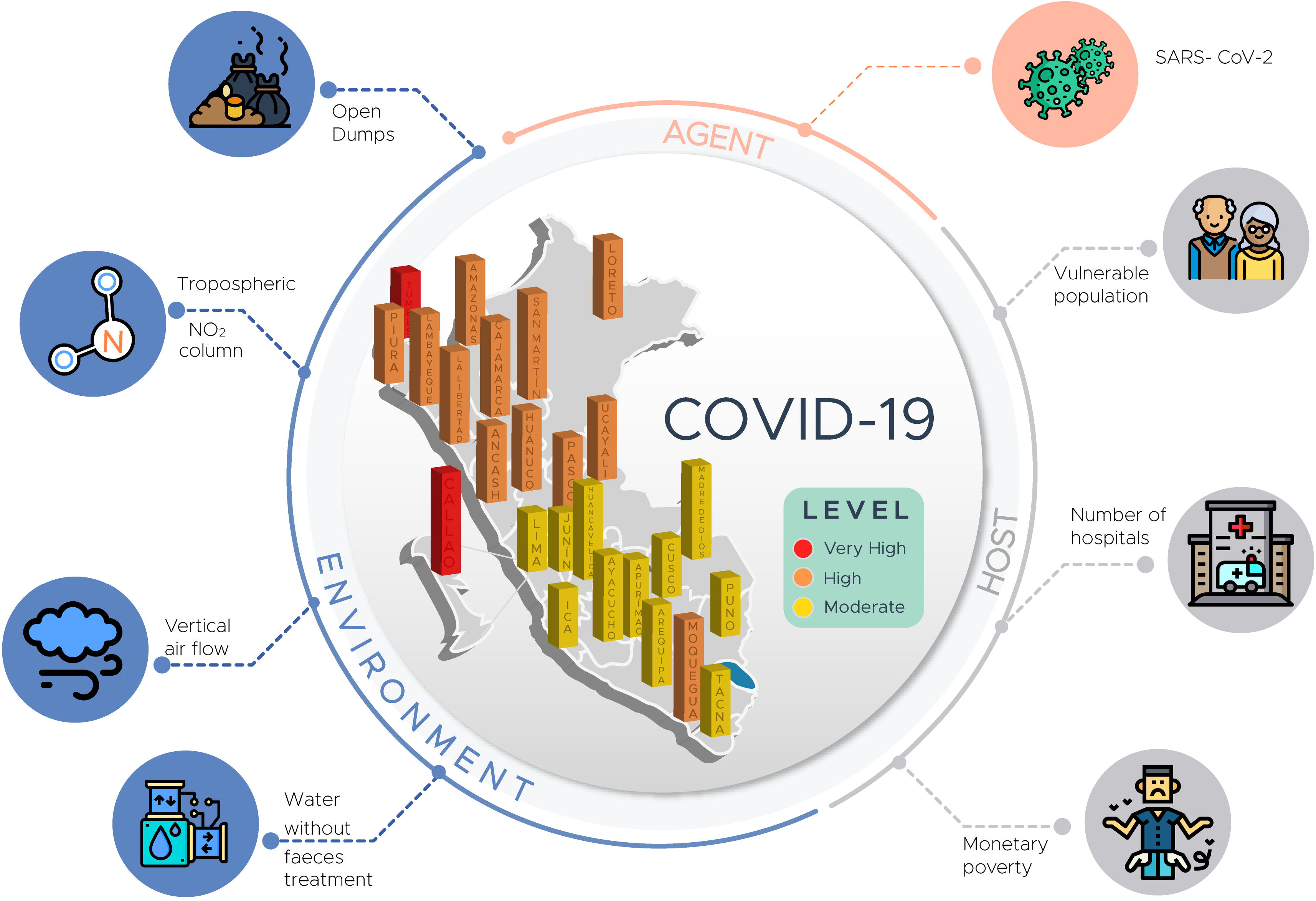

